# Admixture mapping screening of CKD traits and risk factors in U.S. Hispanic/Latino individuals from Central America country-of-origin

**DOI:** 10.1101/2022.06.17.22276554

**Authors:** Andrea R.V.R. Horimoto, Quan Sun, James P. Lash, Martha L. Daviglus, Jianwen Cai, Karin Haack, Shelley A Cole, Timothy A. Thornton, Sharon R. Browning, Nora Franceschini

## Abstract

Chronic kidney disease (CKD) is prevalent in Central America and ancestry-specific factors may contribute to CKD risk. To understand the genetic ancestry influences on CKD susceptibility, we conducted an admixture mapping of CKD traits and risk factors on 1,023 participants of the Hispanic Community Health Study/Study of Latinos who reported four-grandparents originating from the same Central America country. Admixture mapping signals were validated on 8,191 African Americans from the Women’s Health Initiative, 3,141 American Indians from the Strong Heart Study and 197,272 White individuals from the Million Veterans Program. We identified five novel ancestry-derived loci on chromosomes: 14 for albuminuria; 2, 6 and 9 for CKD; and 3 for type 2 diabetes (T2D). The 14q24.2 locus was validated in American Indians and consisted of two regions spanning the *RGS6* gene, in which the European (risk) and Native American (protective) ancestries had opposite effects for albuminuria. This locus was also identified using the traditional association mapping. Among the three CKD loci, the 6q25.3 African ancestry-derived locus at *ARID1B* gene, associated with increased risk for CKD, was validated in African Americans. The T2D locus at 3q22.2 encompasses the *EPHB1* and *KY* genes and was validated in White individuals. U.S. Hispanic/Latino populations are culturally and genetically diverse. Our strategy of using grandparent country-of-origin for selection of a more genetically similar group likely helped the gene discovery. This study of CKD traits and risk factors in individuals from Central America country-of-origin provides new insights into the ancestry-of-origin influences on CKD in this population.

## Introduction

Chronic kidney disease (CKD) is highly prevalent in Central America in countries such as Nicaragua, Costa Rica, El Salvador and Guatemala, and a main cause of death. The estimated overall age-standardized mortality rate of CKD in the region has increased by 60.9% from 1990 to 2017 ^1^. The high burden of CKD combined with limited resources has had a devastating impact in communities. Limited data are available for CKD causes in most of Central America. Several small studies suggest a role of occupational exposures and possibly heat stress for CKD of unknown etiology (CKDu), a disease affecting farming communities in lowland Central America Pacific regions ^2,3^. The genetic susceptibility of CKD in Central America has been understudied.

Understanding the genetic architecture of this population could help to uncover their genetic risk to chronic diseases such as CKD. It could also inform gene-environment interaction studies of toxic exposures that promote disease risk in the context of genetic susceptibility, as has been proposed for CKDu ^2^. A main issue in studying genetic risk factors for CKD in Central America is understanding the ancestry admixture of the population, given genetic risk can be driven by this admixture and population-specific genetic variants in less characterized populations such as Native Americans. We have previously shown that U.S. Hispanic and Latino individuals with ancestry from Mexico, Central America and South America have a high proportion of Native American ancestry in the Hispanic Community Health Study/Study of Latinos (HCHS/SOL)^4^ and that ancestry-related genomic regions are associated with CKD traits ^5,6^. A recent study compared the HLA allele and haplotype frequencies in Amerindians of Costa Rica and Nicaragua ^7^. However, to our knowledge, previous studies did not compare the genetic background of Hispanic/Latino populations from different countries within Central America in relation to CKD risk. HCHS/SOL participants include first- and second-generation U.S. immigrants, and 1,023 participants reported four-grandparents originating from the same Central America country. We used data from these participants to examine the effect of ancestry on CKD susceptibility in Central America. Here, ancestry represents the subjects’ genetic background but not race or ethnicity, which are two social constructs as discussed elsewhere ^8^. We performed an admixture mapping screening to identify ancestry-specific genomic regions associated with CKD traits and CKD clinical risk factors.

## Material and methods

### Population and phenotypes

The HCHS/SOL is a population-based longitudinal cohort study of 16,415 self-identified Hispanic or Latino persons aged 18-74 years recruited from households in predefined census-block groups from four US field centers (Chicago, Miami, the Bronx, and San Diego) between 2008 and 2011 as previously described ^9^. Participants self-identified Hispanic/Latino backgrounds were Central American (n = 1,730), Cuban (n = 2,348), Dominican (n = 1,460), Mexican (n = 6,471), Puerto Rican (n = 2,728), and South American (n = 1,068). HCHS/SOL participants underwent a baseline clinical examination including biological, behavioral, and sociodemographic assessments. Participants provided a random urine specimen and a fasting blood specimen. Urine albumin (mg/dl) and urine/serum creatinine (g/dl) were measured using an immunoturbidometric method and a creatinase enzymatic method traceable to an isotope dilution mass spectrometry (IDMS) reference method, respectively. Estimated glomerular filtration rate (eGFR) was calculated using the Chronic Kidney Disease Epidemiology Collaboration serum creatinine-based equation ^10^. CKD was defined by an untransformed eGFR < 60 ml/min/1.73 m^2^. Albuminuria was defined by an albumin to creatinine ratio (ACR) ≥ 30 mg/g. eGFR and ACR were inverse normal transformed to satisfy the normality assumptions. Type 2 diabetes (T2D) was defined by a fasting glucose >126 mg/dL and/or use of diabetic medications, and hypertension was defined by a blood pressure >140/90 mmHg or use of blood pressure lowering medications. HCHS/SOL protocols were approved by the institutional review boards at each field center, and all subjects provided written informed consent. This study included 1,023 participants who: (a) had all four grandparents’ origin from the same Central America country, based on reported information; (b) were assigned to the Central America genetic-analysis group ^4^; and (c) have given informed consent. The number of subjects included in each analysis varied according to the data availability: albuminuria and ACR (n= 955), CKD and GFR (n = 1,021), and diabetes and hypertension (n = 1,023).

### Genotyping and local ancestry calls

Participants were genotyped in over 2.5 million single nucleotide polymorphisms (SNPs) using a customized Illumina array (HumanOmni2.5-8v1-1), and imputation was performed using the 1000 Genomes Project phase I reference panel. The quality control procedures and details about genotyping, imputation and estimation of local ancestry calls were previously described ^4,11,12^. Principal components (PCs) were estimated in an unrelated subset of HCHS/SOL subjects, excluding 19 subjects with substantial Asian ancestry ^4^. We used the HCHS/SOL genotype data and reference samples from the Human Genome Diversity Project, HapMap phase 3 and 1000 Genomes project Consortium to estimate European, African and Native American ancestries at genomic locations ^12^. Global ancestry proportions were estimated averaging the local ancestry calls across the genome.

### Statistical analyses

Admixture mapping is implemented using a mixed model framework with a multivariate test of European, African, and Native American ancestries jointly in the GENESIS R package ^13^. We first fit models under the null hypothesis of no genetic effect, including random (pairwise kinship coefficients, household, and census block group) and fixed effects (first five principal components, recruitment center [except for CKD], age, sex, sampling weights, and country background). We removed recruitment center from the CKD analysis because the model was not able to converge due to a missing category. Here, country background reflects the common origin of the four grandparents and was included to account for non-genetic differences among subjects. We then used the null models to test for the association between the local ancestry at each locus and the outcomes using a score test. A lead SNP representing the local ancestry at each locus was included in the analyses, for a total of 15,500 studied loci. For each locus that showed significant findings in the joint test, we tested each ancestry against the others to identify the ancestry accounting for the association. Statistical details about the admixture mapping logistic and linear mixed models are discussed elsewhere ^6^. Here, a locus is defined as a long-range admixture-linkage disequilibrium (LD) block created by recent admixture in Hispanic/Latino populations, and local ancestry as the locus-specific ancestry allelic dosages (0, 1 or 2 copies of European, African, or Native American alleles) estimated from the genotype data. A significance threshold was considered 5.7×10^−5^ to account for multiple testing, as defined previously ^14^. Effect sizes were estimated using the allelic dosage of the ancestry driving the signal.

The genome-wide association study (GWAS) was performed using GENESIS R package ^13^ on imputed SNPs from the 1000 Genome Project reference, which were filtered based on IMPUTE2 ^15^ info > 0.8, ratio of observed variance of imputed dosages to the expected binomial variance (> 0.8), and the effective minor allele count > 30 for quantitative and >50 for binary traits. We used the null models previously fit to test the association between each SNP (reference allele dosage) and the outcomes using a score test. It is important to note that both admixture mapping and GWAS were conducted using similar statistical models to avoid bias due to adjustments.

### Loci fine-mapping

Conditional admixture mapping analyses were conducted by including the ancestry-derived allele dosage of the lead (local ancestry) SNPs as covariates to provide further evidence of their implication with the observed association signals. For albuminuria, we performed an additional conditional analysis including an adjustment for the reference allele dosage of the GWAS associated SNP on the original model to investigate if admixture mapping and GWAS associations were tagging the same underlying haplotype.

Variants and genes under the associated loci were annotated using Variant Effect Predictor ^16^, Ensembl ^17^, Combined Annotation Dependent Depletion (CADD) scaled score (cutoff > 10 for potential deleteriousness) ^18^, GWAS catalog ^19^, GeneCards ^20^, and FORGE2. We used ROADMAP Epigenetic data in FORGE2 to identify tissue- and type-specific variants overlapping with epigenetic elements (DNase I hypersensitive sites [DHSs], histone mark chromatin immunoprecipitation broadpeaks, and hidden Markov model chromatin states).

### Validation of findings

We validated our admixture mapping findings using admixture mapping in the Women’s Health Initiative (WHI) ^21^, and association mapping results from the Strong Heart Study (SHS) ^22^ and the Million Veterans Program (MVP) ^23^. Local ancestry calls were not available to conduct admixture mapping in the SHS and MVP datasets. Briefly, 8,191 WHI African American samples had local ancestry calls estimated as previously reported ^24^ and were used to validate the CKD chromosomes 2 and 6 associations, which were driven by African ancestry. Admixture mapping for CKD, defined as described above, was conducted within 2q37.1 and 6q25.3 regions of interest applying mixed models adjusted by age and top four PCs and using phenotype and approaches as previously defined for HCHS/SOL.

We used genome-wide genetic data from 3,141 American Indian participants from SHS to validate genomic regions identified for albuminuria (chromosome 14), CKD (chromosome 9) and T2D (chromosome 3), which showed Native American ancestry-derived associations. We performed SNP association analyses at selected regions, defined by the boundaries of each associated admixture mapping block. Models were adjusted for age, sex, recruitment center and 10 PCs within a mixed model framework using the Efficient and Parallelizable Association Container Toolbox (EPACTS) pipeline. Briefly, the SHS is the largest, well-characterized cohort of U.S. American Indians ^22^. CKD and albuminuria were defined using the same eGFR equation and threshold described for HCHS/SOL. The SHS protocols were approved by the Indian Health Services IRB, IRBs of all Institutions and by the participating Tribal Review Boards. A total of 3,221 participants were selected for genotyping based on available DNA at visit 1. Genome-wide genotyping was performed using the Infinium Multi-Ethnic Global-8 array (MEGA) for 1,748,250 markers. Quality control was performed by excluding samples with a call rate < 0.95, duplicates (n=22) and sex-discordant samples. Variants were excluded if they did not map to chromosome positions or had more than 0.05 missing genotype data. We computed the genetic relatedness matrix and PCs using PLINK 1.9 ^25^. Imputation used the TOPMed freeze 8 reference panel ^26^, after removing SNPs with a missing rate ≥ 10%. About 14 million post-imputation variants with an imputation R^2^ ≥ 0.3 were used in the analyses.

Lastly, to validate the European ancestry-derived locus associated with T2D, we used summary statistics for T2D from the 197,272 MVP White individuals available at dbGap. Details on the analysis were described elsewhere ^27^.

*P* values were corrected for false discovery rate (FDR) in all validation analyses, using the p.adjust R function set up for the Benjamim & Hochberg method ^28^.

## Results

The descriptive characteristics and estimated global ancestries for 1,023 HCHS/SOL Central America country-of-origin participants are shown in Table 1. The mean age was 44.5 years, 59.0% were female, 16.5% had diabetes and 25.1% had hypertension. The mean eGFR was 98.6 ± 17.7 ml/min/1.73 m^2^ and the median ACR was 6.6 mg/g. The overall prevalence of CKD based on low eGFR was about 2.0% and increased albuminuria varies from 9 to 13%. The average global ancestry proportions were 0.46 Native American and 0.13 African but varied by country of grandparent origin. There was a higher Native America proportion among participants reporting grandparents of origin from Guatemala and El Salvador compared to other Central American countries, and a higher African ancestry proportion among those from Honduras compared with other countries-of-origin.

**Table1.**
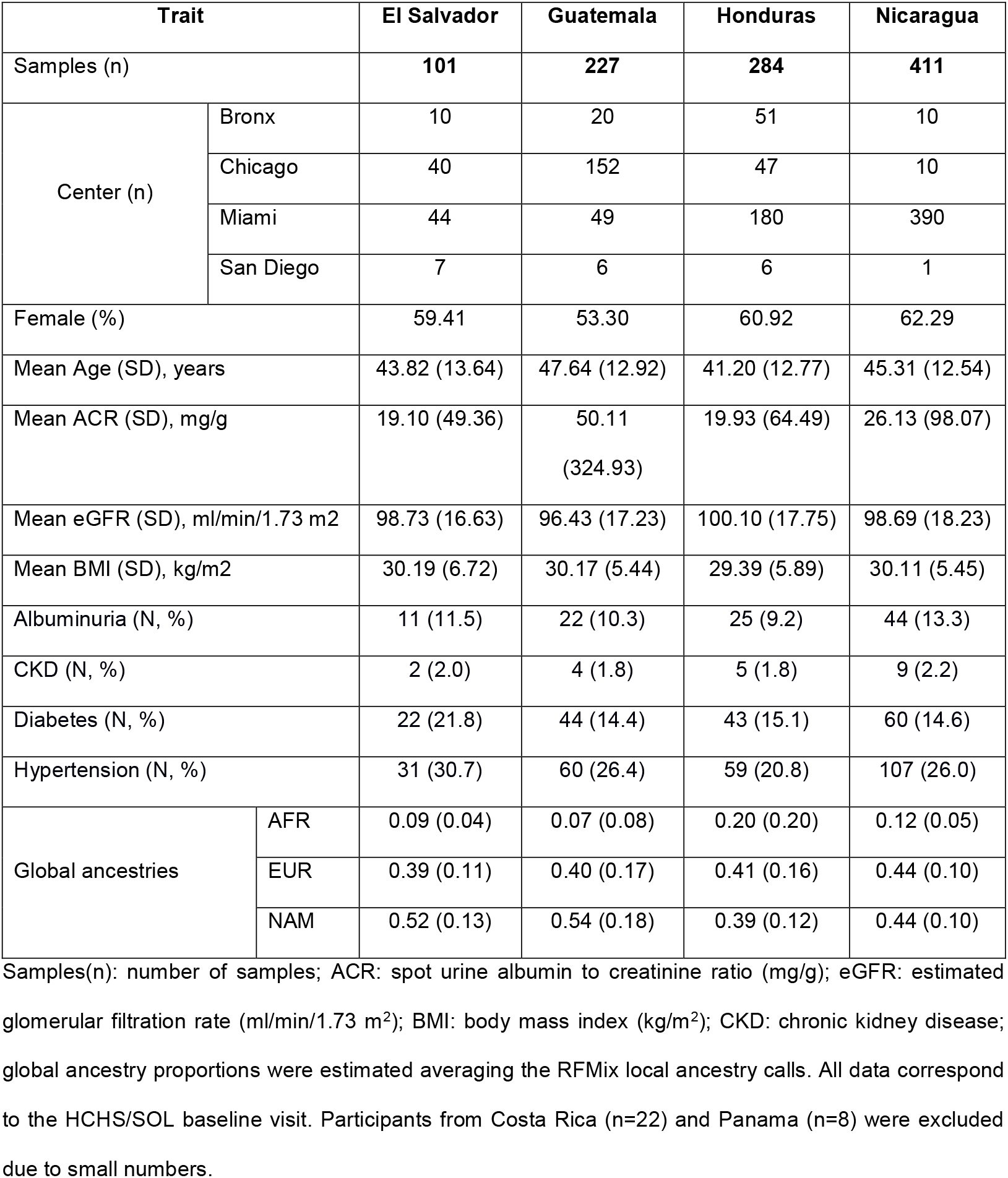
Descriptive characteristics of HCHS/SOL Central American samples.

### Admixture mapping findings in HCHS/SOL

We observed a significant effect of country-of-origin on hypertension. Participants whose grandparents were originated from Honduras (OR: 0.50, 95%CI = 0.25 - 0.97; *P* = 0.04) and Nicaragua (OR: 0.46, 95%CI = 0.23 -0.92; *P* = 0.03) had lower risk of hypertension compared with individuals with ancestry from El Salvador in adjusted models.

We identified two admixture mapping loci for albuminuria at 14q24.2 (122kb and 30kb, respectively, *P* = 3.99×10^−7^; **Fig 1A**), spanning the protein-coded gene *RGS6*. The associations were driven by European and Native American local ancestries, which conferred opposite effects on albuminuria (**Fig 2A**; **Table 2**). While the European-derived allele provided a risk effect (OR: 1.74; *P* = 7.82×10^−4^), the risk for albuminuria decreased for each copy of the Native American allele at this region (OR: 0.44; *P* = 2.85 × 10^−6^).

**Figure 1.**
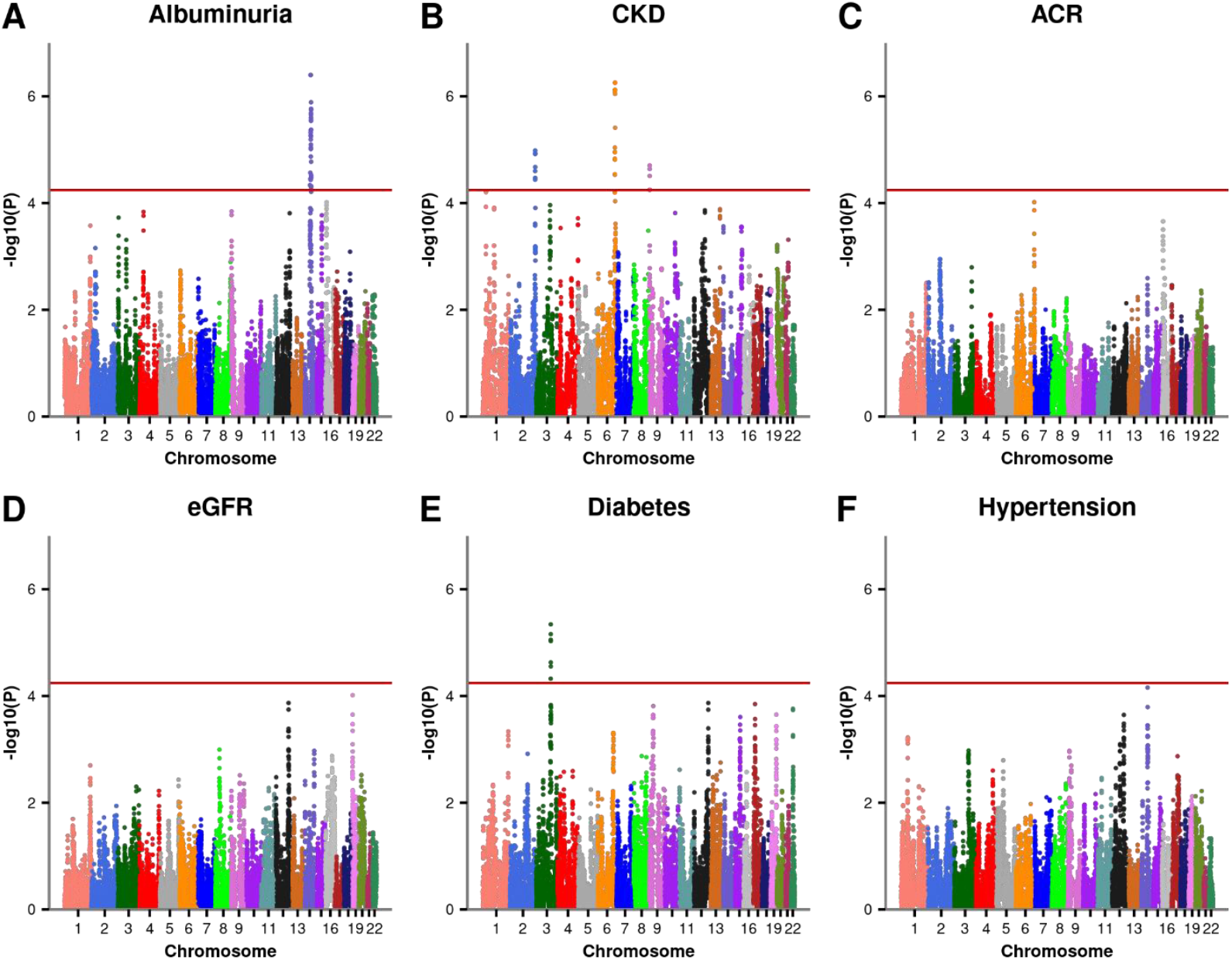
Admixture mapping for CKD-related traits (A to D) and CKD risk factors (E and F) in HCHS/SOL Central American samples.

**Figure 2.**
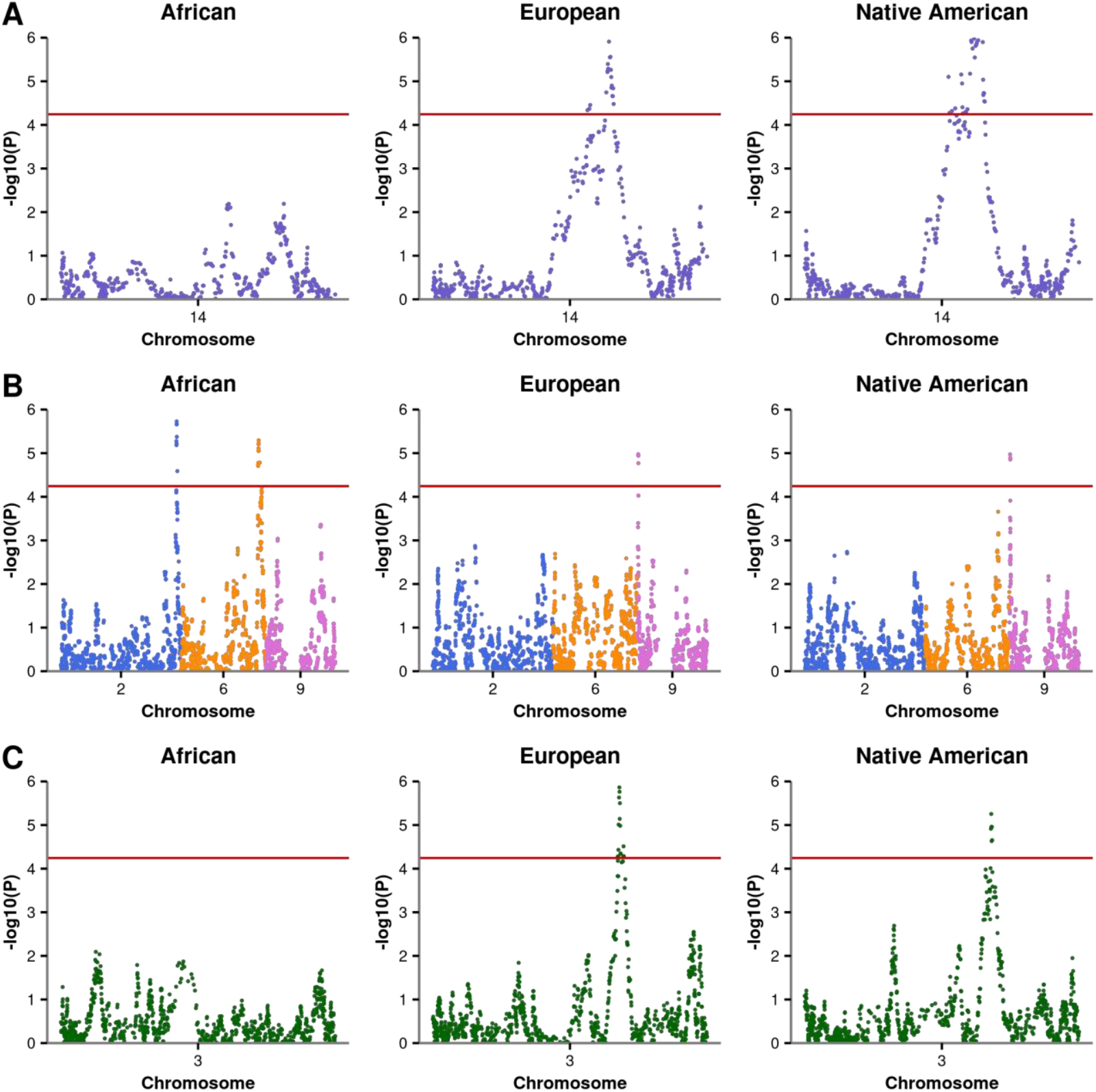
Single ancestry admixture mapping analyses for (A) albuminuria, (B) CKD, and (C) diabetes, in which African, European, and Native American ancestries are tested individually against others. The albuminuria signal on chromosome 14 (A), the CKD signal on chromosome 9 (B), and the diabetes signal on chromosome 3 (C) are driven by both European and Native American ancestries, while the African ancestry is driving the CKD signals on chromosomes 2 and 6 (B).

**Table 2.**
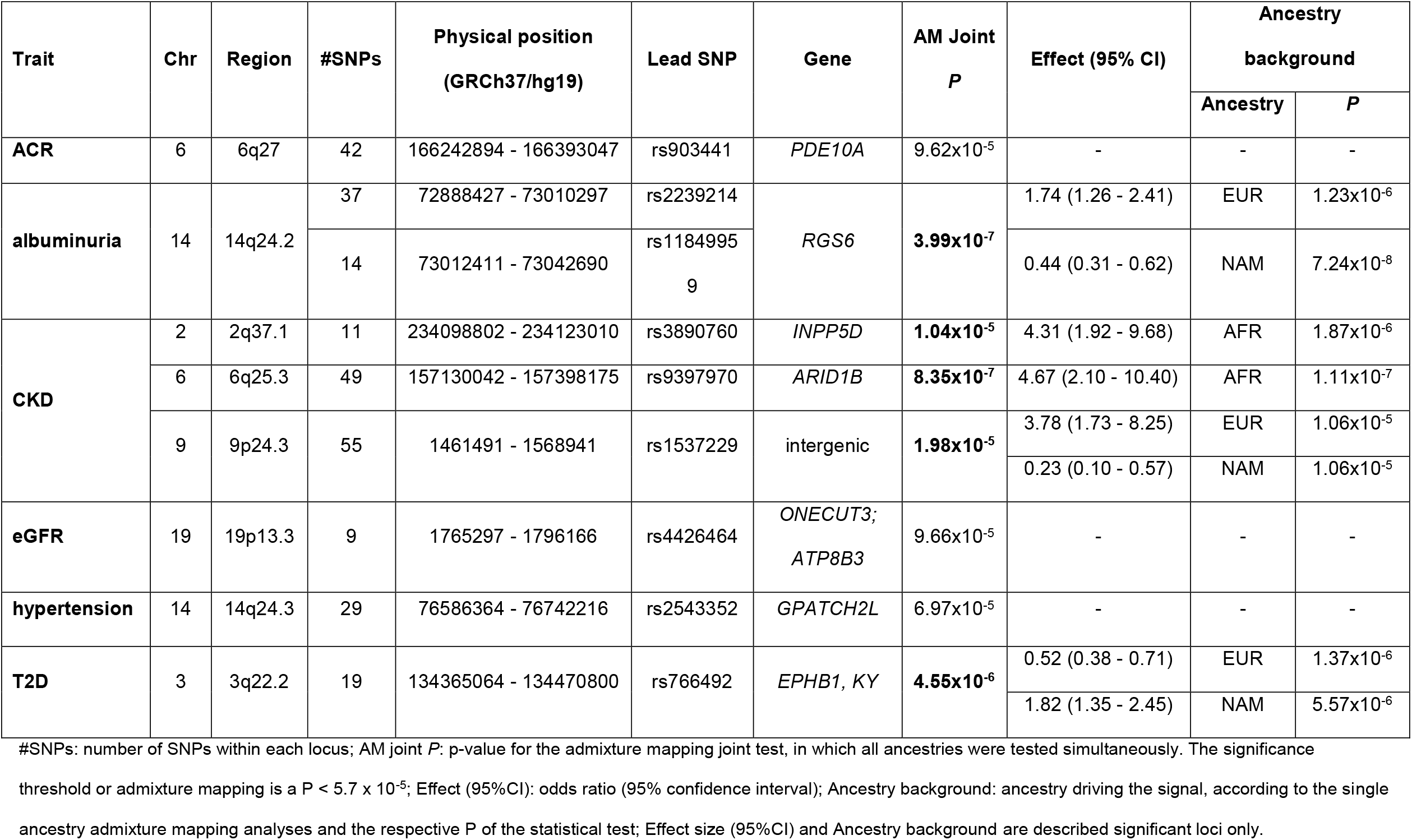
Admixture mapping findings for CKD-related traits and risk factors in HCHS/SOL Central Americans.

| Trait | Chr | Region | #SNPs | Physical position<br>(GRCh37/hg19) | Lead SNP | Gene | AM Joint<br>*P* | Effect (95% CI) | Ancestry<br>background |  |
| --- | --- | --- | --- | --- | --- | --- | --- | --- | --- | --- |
|  |  |  |  |  |  |  |  |  | Ancestry | *P* |
| **ACR** | 6 | 6q27 | 42 | 166242894 - 166393047 | rs903441 | *PDE10A* | 9.62x10<sup>-5</sup> | - | - | - |
| **albuminuria** | 14 | 14q24.2 | 37 | 72888427 - 73010297 | rs2239214 | *RGS6* | **3.99x10<sup>-7</sup>** | 1.74 (1.26 - 2.41) | EUR | 1.23x10<sup>-6</sup> |
|  |  |  | 14 | 73012411 - 73042690 | rs1184995<br>9 |  |  | 0.44 (0.31 - 0.62) | NAM | 7.24x10<sup>-8</sup> |
| **CKD** | 2 | 2q37.1 | 11 | 234098802 - 234123010 | rs3890760 | *INPP5D* | **1.04x10<sup>-5</sup>** | 4.31 (1.92 - 9.68) | AFR | 1.87x10<sup>-6</sup> |
|  | 6 | 6q25.3 | 49 | 157130042 - 157398175 | rs9397970 | *ARID1B* | **8.35x10<sup>-7</sup>** | 4.67 (2.10 - 10.40) | AFR | 1.11x10<sup>-7</sup> |
|  | 9 | 9p24.3 | 55 | 1461491 - 1568941 | rs1537229 | intergenic | **1.98x10<sup>-5</sup>** | 3.78 (1.73 - 8.25) | EUR | 1.06x10<sup>-5</sup> |
|  |  |  |  |  |  |  |  | 0.23 (0.10 - 0.57) | NAM | 1.06x10<sup>-5</sup> |
| **eGFR** | 19 | 19p13.3 | 9 | 1765297 - 1796166 | rs4426464 | *ONECUT3*;<br>*ATP8B3* | 9.66x10<sup>-5</sup> | - | - | - |
| **hypertension** | 14 | 14q24.3 | 29 | 76586364 - 76742216 | rs2543352 | *GPATCH2L* | 6.97x10<sup>-5</sup> | - | - | - |
| **T2D** | 3 | 3q22.2 | 19 | 134365064 - 134470800 | rs766492 | *EPHB1, KY* | **4.55x10<sup>-6</sup>** | 0.52 (0.38 - 0.71) | EUR | 1.37x10<sup>-6</sup> |
|  |  |  |  |  |  |  |  | 1.82 (1.35 - 2.45) | NAM | 5.57x10<sup>-6</sup> |

#SNPs: number of SNPs within each locus; AM joint *P*: p-value for the admixture mapping joint test, in which all ancestries were tested simultaneously. The significance threshold or admixture mapping is a $P < 5.7 \times 10^{-5}$ ; Effect (95%CI): odds ratio (95% confidence interval); Ancestry background: ancestry driving the signal, according to the single ancestry admixture mapping analyses and the respective *P* of the statistical test; Effect size (95%CI) and Ancestry background are described significant loci only.

Admixture mapping of CKD identified three ancestry-derived loci on chromosomes 2, 6, and 9 (**Fig 1B; Table 2**). The chromosome 2 association was captured at 2q37.1 by a 24kb locus comprising 11 SNPs (*P* = 1.04×10^−5^) at *INPP5D* and *ATG16L1* genes, and was driven by African ancestry, which conferred increased risk for CKD (OR: 4.31 for each copy of the African ancestry allele, *P* = 4.09×10^−4^). A similar risk effect of African-derived alleles (OR: 4.67; 1.62×10^−4^) was observed in an associated 268kb region on chromosome 6 which included 49 intronic SNPs of *ARID1B* (6q25.3; 8.35×10^−7^). On chromosome 9, the association was observed in a 107kb intergenic region (9p24.3; P = 1.98×10^−5^) and was driven by both European (OR: 3.78, *P* = 8.68×10^−4^) and Native American (OR: 0.23, *P* = 1.36×10^−3^) ancestries (**Fig 2B; Table 2**), noting that Native American ancestry was protective for CKD.

The admixture mapping for T2D identified an associated locus at 3q22.2 (*P* = 4.55×10^−6^), including 19 SNPs spanning a 106kb region at *KY* and *EPHB1* genes (**Fig 1E; Table2**), which was driven by both European and Native American ancestries (**Fig 2C**). While the European-derived alleles in this region provided a protective additive effect on diabetes (OR: 0.52, *P* = 3.26×10^−5^), the Native American ancestry conferred increased risk of diabetes (OR: 1.82, *P* = 9.21×10^−5^).

Suggestive associations were detected in the admixture mapping for ACR, eGFR, and hypertension (**Fig 1C, 1D, 1F, Table 2**). An association for hypertension at 14q24.3 (*P* = 6.97×10^−5^) was in the same locus identified for albuminuria (*P* = 1.71×10^−6^). The direction of effects was concordant to what we described for albuminuria with an increased risk for hypertension in European ancestry (OR: 1.66, 95%CI = 1.29 - 2.14; *P* = 8.73×10^−5^) and a protective risk in Native American ancestry (OR: 0.62, 95%CI = 0.49 - 0.79; *P* = 1.22×10^−4^).

### SNP association findings in HCHS/SOL and conditional admixture mapping

Association mapping identified a significant variant for albuminuria within 14q24.2 region (**Fig 3A**; rs12885387, 72975499Mb, 4.43×10^−8^). The rs12885387 is an intronic variant of *RGS6*. There were no significant SNP associations at the chromosomes 2, 6 and 9 regions of interest for CKD (**Fig 3B**) and at the chromosome 3 region of interest for diabetes (**Fig S5B**), as well as, within admixture mapping loci with suggestive associations with ACR, eGFR, and hypertension. (**Fig 3C**; **Fig S5A**,**C**).

**Figure 3.**
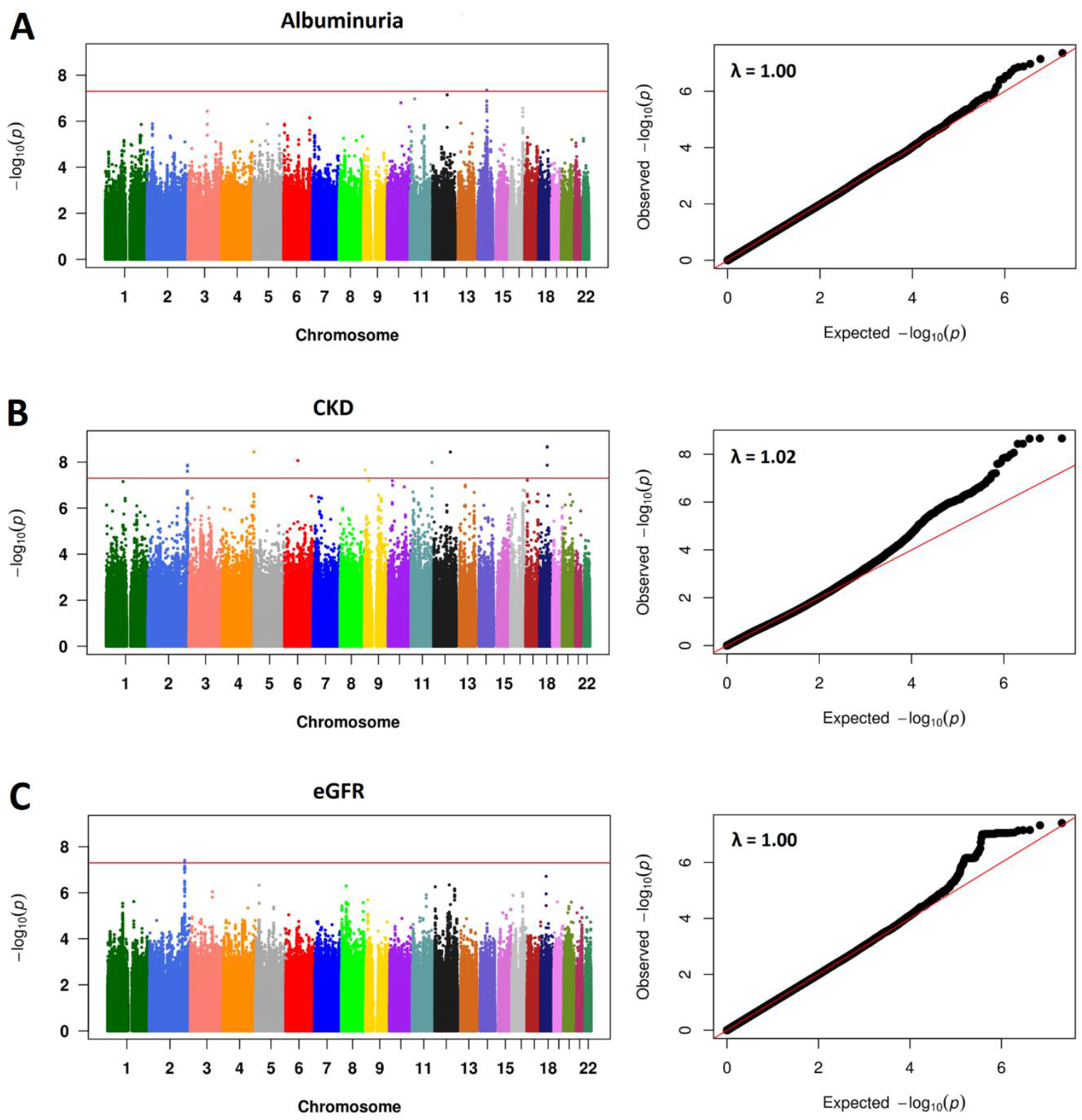
GWAS significant associations identified for CKD-related traits in HCHS/SOL Central American samples. (A) Albuminuria, (B) CKD, and (C) eGFR. Note that significant associations for CKD chromosomes 2, 6, and 9 were observed in different regions of those captured by admixture mapping.

We next performed conditional admixture mapping analyses using local ancestry as a covariate. For regions in which the association was driven by both European and Native American ancestries (3q22.2, 9p24.3 and 14q24.2), we performed the conditional analysis including the ancestry-derived dosages at the locus as covariate (see methods). All association signals for albuminuria (**Fig S1**), CKD (**Fig S2**), and diabetes (**Fig S3**) were not observed in conditional analysis, confirming the role of the ancestry-driven associations at the regions. For albuminuria, we also performed conditional admixture mapping at 14q24.2 with adjustment for the variant rs12885387, which was identified in the association analyses. This SNP explained the admixture association signal (**Fig S4**), suggesting that both admixture mapping and SNP association approaches were tagging the same underlying haplotype.

### Validation of findings

We conducted admixture mapping in 8,191 WHI African Americans for validation of the 2q37.1 and 6q25.3 CKD loci, which was driven by African ancestry. No significant association was observed at 2q37.1 (*P* = 0.96). At the 6q25.3 locus, a region including 70 variants was nominally associated with CKD (*P* = 0.02). The African-derived alleles in this region showed same direction, but lower effect size in WHI (OR: 1.24, 95%CI: 1.03 - 1.49, P = 0.02) compared to HCHS/SOL (OR: 4.67, 95%CI: 2.10 - 10.40, P = 1.62×10^−4^) (**Tables 2 and 3**).

**Table 3.** Validation analyses of associated loci in WHI African American, SHS American Indian, and MVP European samples.

| Trait | Chr | Region | Validation samples | #SNPs | Lead SNP | Physical position (GRCh37/hg19) | Gene | *P* | Effect (95% CI) |
| --- | --- | --- | --- | --- | --- | --- | --- | --- | --- |
| **albuminuria** | 14 | 14q24.2 | SHS | 35 | rs2302144 | 72921417 | *RGS6* | 9.4x10<sup>-5</sup> | 1.09 (1.05 - 1.12) |
| **CKD** | 2 | 2q37.1 | WHI | 7 | rs10203185 | 234102070-234123010 | - | 0.96 | - |
|  | 6 | 6q25.3 | WHI | 70 | rs9397970 | 157130042-157385621 | *ARID1B* | 0.02 | 1.24 (1.03 - 1.49) |
|  | 9 | 9p24.3 | SHS | 1 | rs10114135 | 1470797 | - | 0.49 | - |
| **T2D** | 3 | 3q22.2 | MVP | 41 | rs6439513 | 134430871 | *EPHB1* | 0.03 | 1.07 (1.03 - 1.10) |

Chr: chromosome; #SNPs: number of associated SNPs within each locus (for WHI, it corresponds to the number of SNPs included in the admixture-LD block); *P*: p-value for the admixture mapping joint test for WHI and p-value for the association analysis for SHS and MVP, after FDR correction; Effect (95%CI): odds ratio (95% confidence interval).

For the Native American-driven loci associated with CKD (9p24.3) and albuminuria (14q24.2), we performed target association in 3,141 SHS American Indian individuals using GWAS data. In total, 35 variants reached the region-wide significance threshold for albuminuria after applying FDR-controlling procedures (*P* < 0.05). The most significant finding was rs2302144 (14:72921417; *P* = 9.4×10^−5^; OR = 1.09, 95%CI = 1.05 - 1.12), an intronic variant of *RGS6* gene (**Table 3**). There was no evidence of association at the 9p24.3 CKD locus.

The European T2D locus at 3q22.2 was validated in GWAS data of 197,272 MVP European samples. Forty-one variants reached the regional-wide significance threshold after FDR correction (*P* < 0.05), in which the rs6439513 (3: 134430871; *P* = 0.03; OR = 1.07, 95%CI = 1.03 - 1.10), an intronic SNP of *EPHB1*, was the lead SNP (**Table 3**).

In summary, we validated three loci (3q22.2 [for T2D], 6q25.3 [for CKD] and 14q24.2 [for albuminuria]), but not 2q37.1 and 9p24.3 for CKD.

## Discussion

This study is the first to leverage the genetic ancestry of admixed individuals based on their grandparents’ country-of-origin in Central America and one of the few genetic studies of individuals from this region. It provides important information on the genetic background for CKD susceptibility in this understudied population, which is a first step to elucidate the genetic causes of high prevalence of CKD in Central America and to design genetic studies that incorporate environment interactions for CKD. We conducted gene mapping of CKD traits and its risk factors using genome-wide admixture mapping to identify ancestry-specific genomic regions associated with these clinical phenotypes. Our main findings were the discovery of five novel ancestry-of-origin loci on chromosome 14 for albuminuria, 2, 6 and 9 for CKD, and 3 for T2D, three of which (3q22.2, 6q25.3 and 14q24.2) were validated in African American, American Indian, and European populations.

Among highlighted findings, the 14q24.2 region was identified for albuminuria using both admixture mapping and SNP association approaches. The European and Native American ancestry-derived loci comprised 51 intronic variants of *RGS6* (regulator of G-protein signaling 6) gene. These loci include several variants that overlap epigenetic elements, and two SNPs predicted deleterious (rs917427 and rs2074647 [CADD = 13]) (**Table S1**). *RGS6* is a member of the R7 subfamily of RGS proteins, which are critical for cellular function and a potential therapeutic target for several complex diseases ^29–31^. The gene is highly expressed in heart ^31^ and has been previously implicated with cardiovascular risk factors ^32–34^, including heart rate ^35^, and C-reactive protein (CRP) levels ^36,37^. CRP is a marker for inflammation associated with progression of CKD ^38,39^. Both admixture mapping and SNP association identified the same locus for this trait and conditional admixture mapping supported the tagging of the same haplotype.

Interestingly, another locus at 14q24.3 had suggestive evidence for association with albuminuria and hypertension, and European and Native American ancestries displayed the same direction of effects for these traits. These findings mirror the evidence from observation studies of increased albumin and hypertension. This locus includes 29 variants enriched for regulatory features and epigenetic elements (**Table S2**). Three intronic SNPs of *GPATCH2L* are predicted deleterious (rs2360982, CADD 19.33; rs3813547, CADD 18.30; rs2543380, CADD 11.08), and rs2360982 and rs3813547 overlapped with DHS, transcription site, enhancer and histone marks in kidney tissue. *GPATCH2L* gene has previously shown suggestive association with blood pressure ^40^.

Admixture mapping identified three associations for CKD on chromosomes 2, 6 and 9, but we were only able to validate the 6q25.3 African-derived locus at *ARID1B* in African American individuals. *ARID1B* encodes the AT-Rich Interactive Domain 1B protein, which is a subunit of a chromatin remodeling complex responsible to modulate the transcription, and plays a crucial role in the stem cell pluripotency and cell differentiation and proliferation ^41^. It is also implicated with the maintenance of pericytes and vascular endothelial cells in the heart, being pivotal for survival ^42^. *ARID1B* is expressed in multiple cell types in healthy and diabetic human kidney tissue in scRNAseq databases. Mutations in this gene have been associated with several developmental disorders and cancer ^41,43^ and variants in the gene were recently identified for congenital heart disease ^44^ and blood pressure traits in African ancestry samples ^45^. Several variants in the 6q25.3 loci are involved in regulatory function and overlap epigenetic elements, including five deleterious protein coding variants (CADD > 10) (**Table S3**). Although chromosomes 2 and 9 loci did not validate in independent samples, both loci span variants with regulation and epigenetic features and several SNPs with potential deleterious effects (CADD > 10), including a missense variant of *INPP5D* gene (rs9247, CADD 22.40) at 2q37.1. The 2q37.1 locus spans two protein coding genes (*INPP5D* and *ATG16L1*), while the 9p24.3 locus falls in an intergenic region (**Table S3**). *ATG16L1* is crucial for autophagy processes and has been previously associated with type 2 diabetes and its medication use ^46^.

The 3q22.2 European and Native American ancestry-of-origin loci associated with T2D spans two protein coding genes (*EPHB1* and *KY*) and 19 variants, several of them overlapping with DHSs and histone marks (**Table S4**). *EPHB1* is an ephrin receptor that mediates multiple developmental processes. In animal experiments, *EPHB1* has been associated with diabetic neuropathy ^47^, and is downregulated in diabetic glomeruli ^48^. *EPHB1* variants were associated with body mass index (BMI) in a trans-ethnic GWAS ^46^, and these traits are risk factors for T2D. KY gene encodes a protein involved in the neuromuscular junction physiology that has also been associated with BMI and other anthropometric traits ^19^.

Overall, this study provides evidence for validated of admixture mapping regions for albuminuria, T2D, and CKD at the chromosome 6, but not for CKD associated loci on chromosomes 2 and 9. The African ancestry at the 6q25.3 CKD locus showed a consistent direction of effect between HCHS/SOL and WHI through admixture mapping validation. For loci validated using association mapping (3q22.2 for T2D and 14q24.2 for albuminuria), the direction of effects was not consistent between discovery and validation samples (**Tables 2 and 3**). This is not unexpected given differences in the genetic ancestry background or admixture in these populations, and the differences in the statistical approaches used for analyses. Unlike GWAS, the admixture mapping captures extended haplotypes of ancestry-derived regions associated with outcomes and has no resolution to point out the variants underlying the association signal which could include one or more common or rare variants. The association mapping in the MVP and SHS can only replicate the tested variants and indirectly those that are in LD with them within genomic regions. Therefore, they cannot account for the complex architecture at these loci. Most importantly, we validated the admixture regions in populations for the same ancestry of those that drove the signals.

This study shows that leveraging the genetic ancestry in a more genetically similar group based on their grandparent country of origin can improve the discovery of genes influencing CKD risk in admixed Hispanic/Latino populations. The genetic diversity and the population structure of the HCHS/SOL samples have been previously discussed ^4,12^. Using PCs, HCHS/SOL participants cluster differently across the three African, European and Native American ancestry components.

Individuals whose grandparents originated from Central America tend to cluster separately, especially for the African and Native American components ^12^. Using Individuals from Central America, we were able to identify ancestry-specific regions implicated with CKD traits and risk factors, which were not identified in studies involving the more heterogeneous whole HCHS/SOL samples ^5,6^.

A limitation of our study is the lack of another Central American sample to validate our findings. Populations under the Hispanic/Latino umbrella are genetically and culturally very diverse and remain underrepresented in genetic studies. We were able to validate some of the ancestry-specific regions using African American, American Indian and European population studies. Differences in the genetic background of the HCHS/SOL Central American participants may contribute to our findings, particularly because the type and allele frequencies of variants in individuals from Central America likely vary compared to other populations. Therefore, admixture mapping can provide additional insights by identifying ancestry-related genomic regions associated with disease.

This is the first genetic study to map the CKD-related traits and risk factors in individuals with ancestry background from Central America, which have a high Native American global ancestry proportion and less known genetic variation. This study provides new information into the ancestry-specific genetic contributions to the CKD risk in Hispanic/Latino populations. Our strategy of using grandparent country-of-origin for a more genetically similar group likely help the gene discovery. Although we cannot assess CKDu directly given the lack of clinical diagnosis in our sample, we speculate that our participants could be used as control samples in studies of CKDu given their similar genetic background.

## Data Availability

The Hispanic Community Health Study / Study of Latinos (HCHS/SOL) data supporting the findings of this study is openly available in the dbGap repository at https://www.ncbi.nlm.nih.gov/gap/, under the accession numbers phs000880.v1.p1 and phs000810.v1.p1. The admixture mapping linear and logistic mixed models are implemented in the GENESIS R package freely available at http://bioconductor.org/packages/release/bioc/html/
GENESIS.html. This project used existing genotype and phenotype data from the Women's Health Initiative, which is available through the dbGap (phs000200.v12.p3). The MVP summary statistics is available through dbgap (phs001672.v7.p1).

## Supplemental data

Supplemental data includes five figures and four tables.

## Declaration of interests

The authors declare no competing interests.

## Acknowledgements

The baseline examination of HCHS/SOL was carried out as a collaborative study supported by contracts from the National Heart, Lung, and Blood Institute (NHLBI) to the University of North Carolina (N01-HC65233), University of Miami (N01-HC65234), Albert Einstein College of Medicine (N01-HC65235), Northwestern University (N01-HC65236), and San Diego State University (N01-HC65237). The following Institutes/Centers/Offices contributed to the first phase of HCHS/SOL through a transfer of funds to the NHLBI: National Institute on Minority Health and Health Disparities, National Institute on Deafness and Other Communication Disorders, National Institute of Dental and Craniofacial Research (NIDCR), National Institute of Diabetes and Digestive and Kidney Diseases, National Institute of Neurological Disorders and Stroke, NIH Institution-Office of Dietary Supplements. The Genetic Analysis Center at Washington University was supported by NHLBI and NIDCR contracts (HHSN268201300005C AM03 and MOD03). Genotyping efforts were supported by NHLBI HSN 26220/20054C, NCATS CTSI grant UL1TR000124, and NIDDK Diabetes Research Center (DRC) grant DK063491. This study is supported by the following grants: National Institutes of Health MD012765, HL140385 and DK117445 to N. Franceschini.

## Web resources

GENESIS R package, http://bioconductor.org/packages/release/bioc/html/GENESIS.html

IMPUTE2, https://mathgen.stats.ox.ac.uk/impute/impute_v2.html

Variant Effect Predictor, https://uswest.ensembl.org/info/docs/tools/vep/index.html

Ensembl, https://uswest.ensembl.org/index.html

Combined Annotation Dependent Depletion score, https://cadd.gs.washington.edu/

GWAS catalog, https://www.ebi.ac.uk/gwas/

GeneCards, https://www.genecards.org/

FORGE2, https://forge2.altiusinstitute.org/

ROADMAP Epigenetic data, https://www.encodeproject.org/

EPACTS, https://genome.sph.umich.edu/wiki/EPACTS

PLINK 1.9, https://www.cog-genomics.org/plink/

## Data and code availability

The Hispanic Community Health Study / Study of Latinos (HCHS/SOL) data supporting the findings of this study is openly available in the dbGap repository at https://www.ncbi.nlm.nih.gov/gap/, under the accession numbers phs000880.v1.p1 and phs000810.v1.p1. The admixture mapping linear and logistic mixed models are implemented in the GENESIS R package freely available at http://bioconductor.org/packages/release/bioc/html/GENESIS.html. This project used existing genotype and phenotype data from the Women’s Health Initiative, which is available through the dbGap (phs000200.v12.p3). The MVP summary statistics is available through dbgap (phs001672.v7.p1).

## Notes

### Competing Interest Statement

The authors have declared no competing interest.

### Author Declarations

The Hispanic Community Health Study / Study of Latinos (HCHS/SOL) data supporting the findings of this study is openly available in the dbGap repository at https://www.ncbi.nlm.nih.gov/gap/, under the accession numbers phs000880.v1.p1 and phs000810.v1.p1. This project used existing genotype and phenotype data from the Women's Health Initiative, which is available through the dbGap (phs000200.v12.p3). The MVP summary statistics is available through dbgap (phs001672.v7.p1).

