## Supplementary material for "Admixture mapping screening of CKD traits and risk factors in U.S. Hispanic/Latino individuals from Central America country-of-origin": Fig S

**Supplemental figures**

**Figure S1**. Conditional admixture mapping analyses for albuminuria including the lead SNPs of the two most associated loci (rs2239214 and rs11849959) as covariates in the model.

**Figure S2**. Conditional admixture mapping analyses for CKD including the lead SNPs of the chromosomes 2 (A), 6 (B), and 9 (C) loci as covariates in the model.

**Figure S3**. Conditional admixture mapping analyses for diabetes including the lead SNP rs766492 (European [left] and Native American [right] dosages) as covariate in the model.

**Figure S4.** Conditional admixture mapping analysis on chromosome 14 for albuminuria including the GWAS associated SNP rs12885387 as covariate.

**Figure S5.** GWAS suggestive associations identified for (A) ACR, (B) diabetes and (C) hypertension in HCHS/SOL Central American samples.

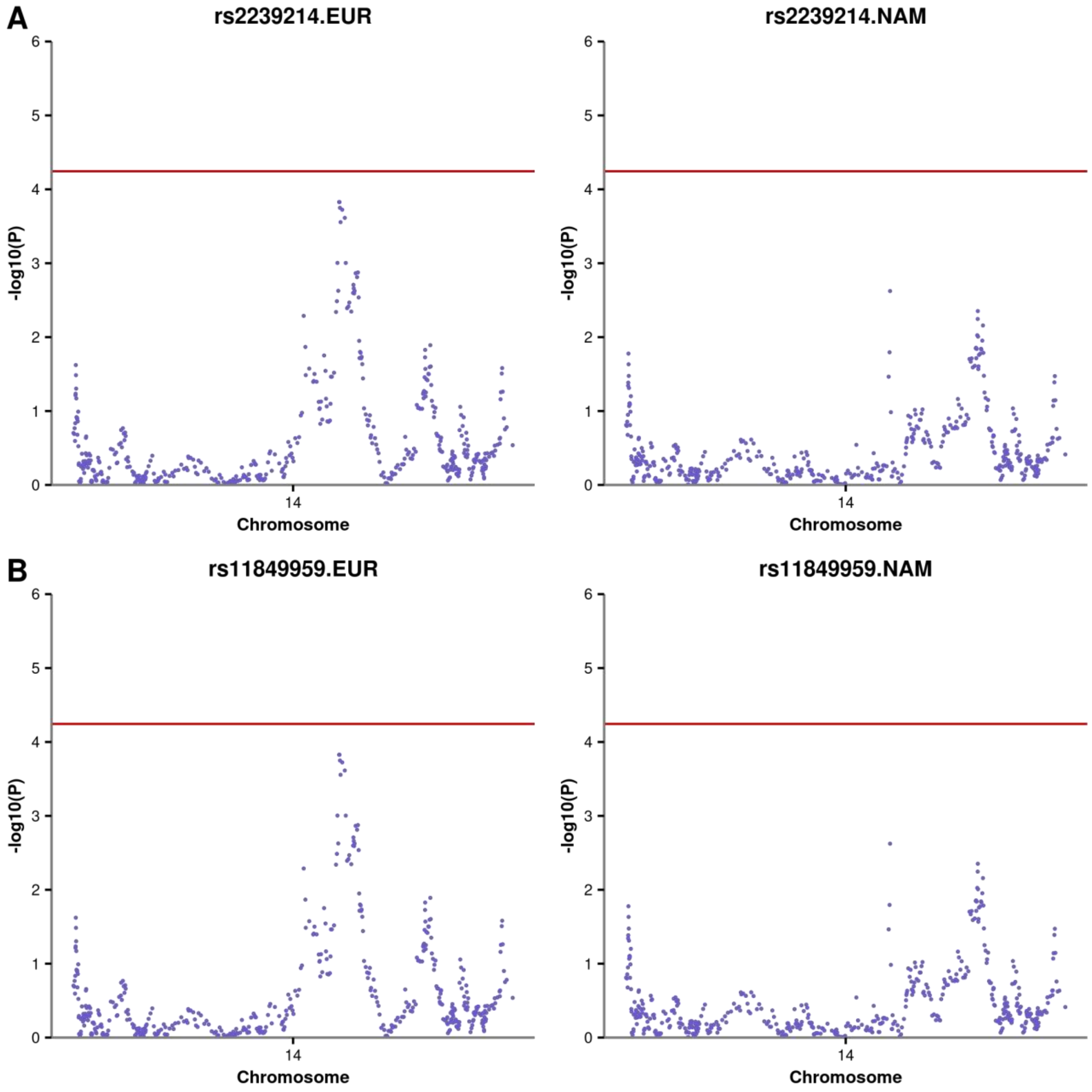


**Figure S1**. **Conditional admixture mapping analyses for albuminuria including the lead SNPs of the two most associated loci (rs2239214 and rs11849959) as covariates in the model.** Note that we conducted the analyses using the European and Native American allelic dosages of each lead SNP because both ancestries were driving the association signal. (A) rs2239214 using European allelic dosages (left) and Native American allelic dosages (right); (B) rs11849959 using European allelic dosages (left) and Native American allelic dosages (right).

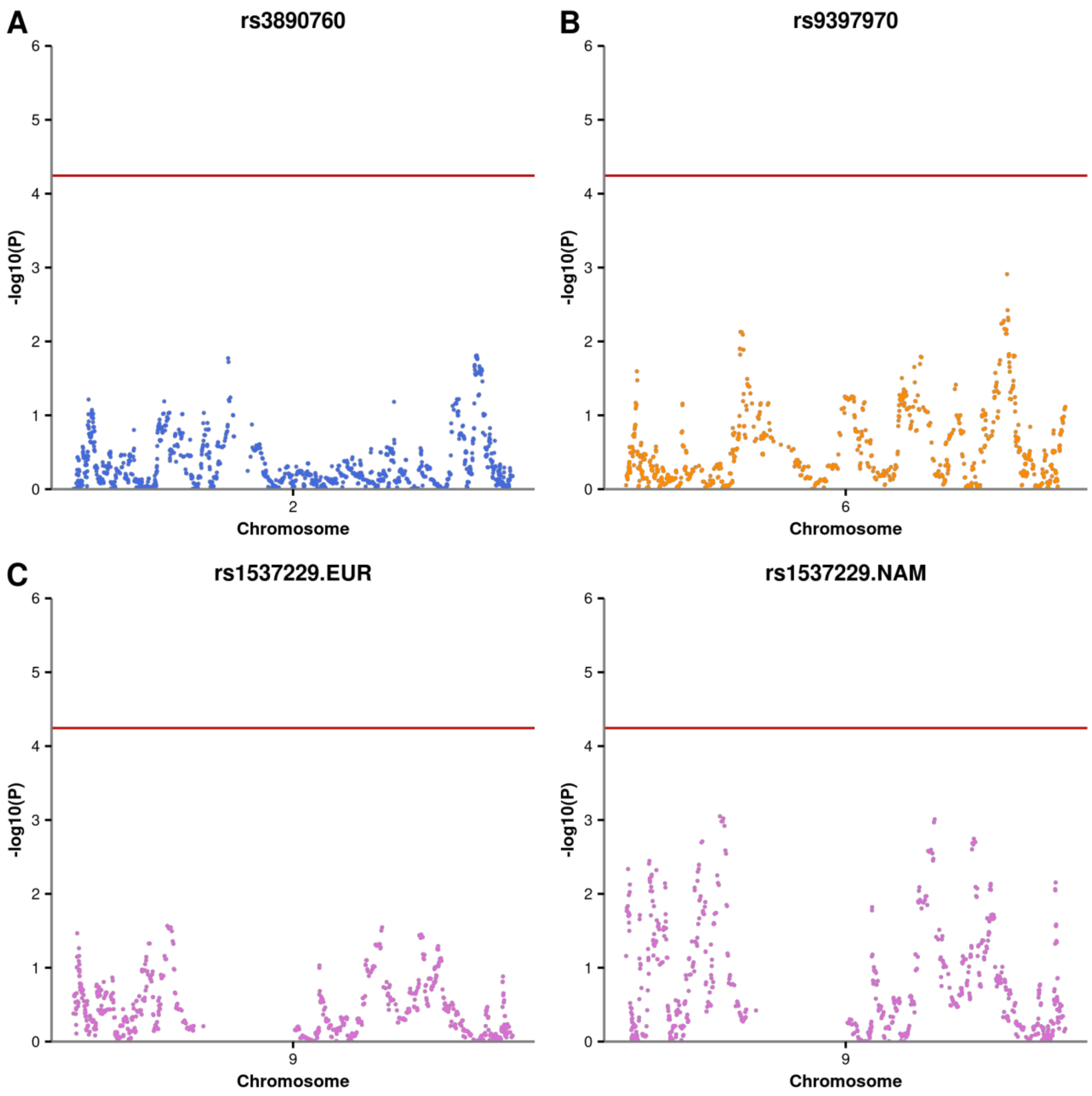


**Figure S2**. **Conditional admixture mapping analyses for CKD including the lead SNPs of the chromosomes 2 (A), 6 (B), and 9 (C) loci as covariates in the model.** The conditional analyses on chromosomes 2 and 6 were performed using the African allelic dosages of the lead SNPs (A) rs3890760 and (B) rs9397970, respectively. For the chromosome 9 analyses, we used the European (left) and Native American (right) allelic dosages of the lead SNP (C) rs1537229.

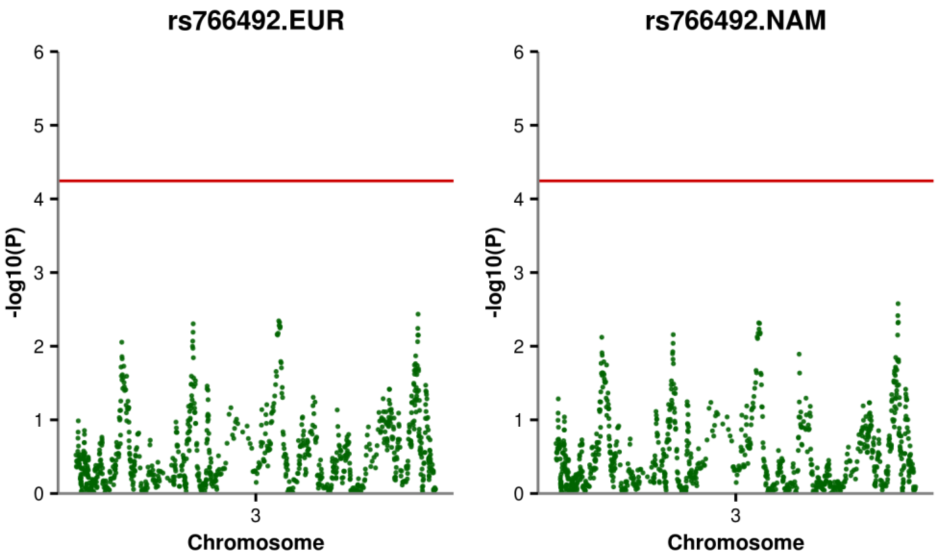


**Figure S3**. **Conditional admixture mapping analyses for diabetes including the lead SNP rs766492 (European [left] and Native American [right] dosages) as covariate in the model.**

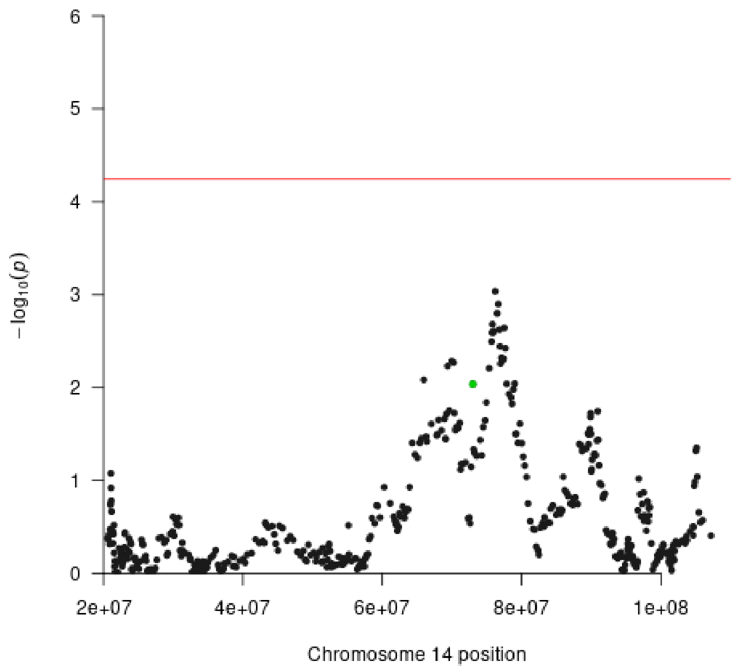


**Figure S4. Conditional admixture mapping analysis on chromosome 14 for albuminuria including the GWAS associated SNP rs12885387 as covariate.** The admixture mapping lead SNPs on 14q24.2 (rs2239214 and rs11849959) are highlighted in green.

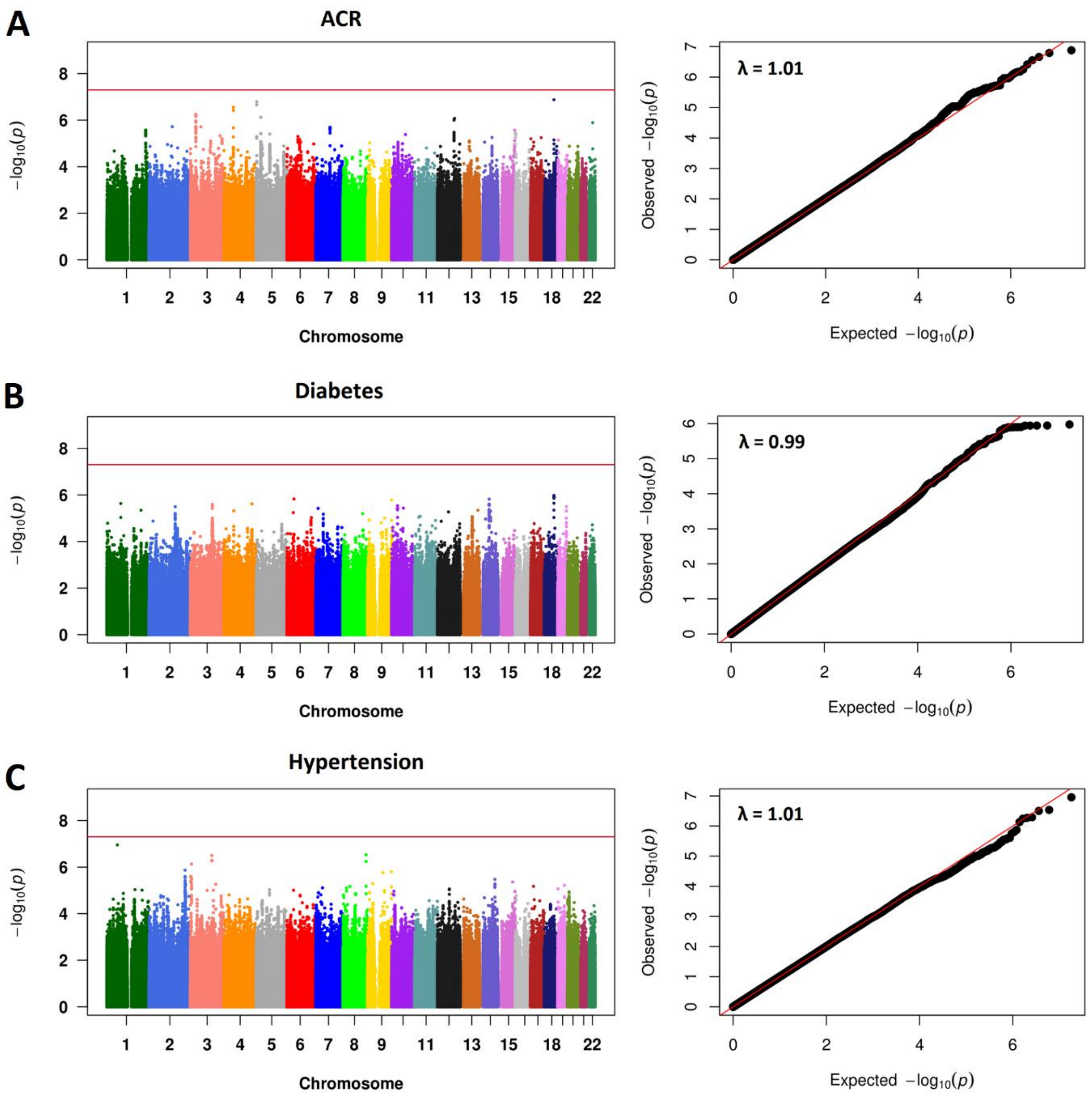


**Figure S5. GWAS suggestive associations identified for (A) ACR, (B) diabetes and (C) hypertension in HCHS/SOL Central American samples.**
